# Functional myeloid-derived suppressor cells expand in blood but not airways of COVID-19 patients and predict disease severity

**DOI:** 10.1101/2020.09.08.20190272

**Authors:** Sara Falck-Jones, Sindhu Vangeti, Meng Yu, Ryan Falck-Jones, Alberto Cagigi, Isabella Badolati, Björn Österberg, Maximilian Julius Lautenbach, Eric Åhlberg, Ang Lin, Inga Szurgot, Klara Lenart, Fredrika Hellgren, Jörgen Sälde, Jan Albert, Niclas Johansson, Max Bell, Karin Loré, Anna Färnert, Anna Smed-Sörensen

## Abstract

The immunopathology of COVID-19 remains enigmatic, exhibiting immunodysregulation and T cell lymphopenia. Monocytic myeloid-derived suppressor cells (M-MDSC) are T cell suppressors that expand in inflammatory conditions, but their role in acute respiratory infections remains unclear. We studied blood and airways of COVID-19 patients across disease severity at multiple timepoints. M-MDSC frequencies were elevated in blood but not in nasopharyngeal or endotracheal aspirates of COVID-19 patients compared to controls. M-MDSCs isolated from COVID-19 patients suppressed T cell proliferation and IFNγ production partly via an arginase-1 (Arg-1) dependent mechanism. Furthermore, patients showed increased Arg-1 and IL-6 plasma levels. COVID-19 patients had fewer T cells, and displayed downregulated expression of the CD3ζ chain. Ordinal regression showed that early M-MDSC frequency predicted subsequent disease severity. In conclusion, M-MDSCs expand in blood of COVID-19 patients, suppress T cells and strongly associate with disease severity, suggesting a role for M-MDSCs in the dysregulated COVID-19 immune response.

## Introduction

The pathogenesis of COVID-19 caused by severe acute respiratory syndrome coronavirus-2 (SARS-CoV-2) remains elusive. SARS-CoV-2 infection ranges from asymptomatic disease to multi-organ failure and death^1^. COVID-19 is characterized by influenza-like symptoms (including fever, cough and myalgia), and in severe cases, respiratory failure and acute respiratory distress syndrome, occurring in around 40% of hospitalized cases^1, 2, 3^. Fatal COVID-19 is caused by tissue-directed immunopathology, especially in the lungs, rather than the virus itself^4, 5^. Furthermore, it is known that immune cells differ depending on their anatomical location^6, 7, 8, 9, 10^. Therefore, studying both systemic and respiratory immune responses in COVID-19 is important to fully understand its pathogenesis and to identify factors dictating disease severity.

COVID-19 is associated with substantial immune activation including elevated levels of proinflammatory cytokines such as IL-6^11^. Furthermore, T cell lymphopenia occurs, especially in critical cases^12^, but the underlying mechanism(s) remain unclear. SARS-CoV-2 specific T cells are important in combating the virus^13^, and a functional T cell response is critical for clearing infections in general. Myeloid-derived suppressor cells (MDSCs) are myeloid immune cells with an immature phenotype and potent T cell suppressive capacity^14, 15, 16^. MDSCs expand in inflammatory conditions including cancer, autoimmune disease and chronic viral infections like HIV and hepatitis C^17^. Two subpopulations of MDSCs have been identified based on phenotypic and morphological features: monocytic MDSCs (M-MDSCs) and polymorphonuclear MDSCs (PMN-MDSCs) with partly overlapping functions^18^. MDSC driven mechanisms of T cell suppression include secretion of arginase 1 (Arg-1) thereby catabolizing L- arginine, generation of reactive oxygen species (ROS) and nitric oxide (NO), direct engagement of T cell inhibitory and apoptotic receptors, and production of inhibitory cytokines such as IL-10 and TGF-β^19^.

Single-cell RNA sequencing, mass cytometry and flow cytometry on blood have suggested that expansion of suppressive myeloid cells is a hallmark of severe COVID-19^20, 21^. Furthermore, high frequency of PMN-MDSCs was recently reported to correlate with disease severity in COVID-19^22^. However, functional and mechanistic data on the role of M-MDSCs during COVID-19 are lacking, further confounded by the lack of knowledge of the role of M-MDSCs in respiratory infections in general.

In this study, we investigated M-MDSCs in COVID-19 patients across disease severity and compared to influenza patients and healthy controls. Influenza A virus infection is often compared to COVID-19 due to similarities including the diverse clinical presentation and the route of transmission^23, 24^. We found a striking association between the frequency of blood M-MDSCs and COVID-19 disease severity. However, the frequency of M-MDSCs from the nasopharynx and lower airways did not correlate with disease severity in COVID-19. Importantly, purified M-MDSCs were functional and suppressed T cell proliferation, partly via an Arg-1 dependent mechanism. In line with this, plasma Arg-1 levels were elevated in COVID-19 patients in a disease severity dependent manner. Finally, we found that early frequency of blood M-MDSCs predicted subsequent disease severity, suggesting both that M-MDSCs are involved in the dysregulation of the immune response in COVID-19 and that they may be used as a potential prognostic marker in COVID-19 patients.

## Results

### Study subject characteristics

In total, 147 adults with PCR confirmed SARS-CoV-2 infection, ranging from mild to fatal disease were enrolled in the study: 91 patients from hospital wards, 43 patients from the ICU, 3 patients from an outpatient clinic and 10 household contacts, and blood as well as respiratory samples were collected longitudinally (Figure 1A and Supplementary figure 1). Identical samples from 44 patients with PCR confirmed influenza A virus infection with mild to moderate disease, as well as from 33 age-matched healthy controls (HCs) were included for comparison (Figure 1B and Table 1). As expected, disease severity in the COVID-19 patient cohort varied over time (Figure 1C, Table 2 and Supplementary figure 1), and some patients deteriorated during their hospital stay. At peak disease severity, 13% of patients were classified as having mild disease, 39% as moderate, and 39% as severe (Figure 1C). Furthermore, there were 12 recorded fatalities (8.1%) in the COVID-19 cohort during the observation period (Figure 1C). The peak disease severity score prior to death was 6 in all but two patients who had scores of 4 and 5 respectively. At the end of the study period, 110 of the non-fatal, hospitalized patients had been discharged while 12 patients remained in hospital all of whom were already classified as having severe disease. The distribution of age varied significantly across peak disease severity groups in COVID-19 patients (p< 0.001), as did body mass index (BMI) (p< 0.001), male gender (p = 0.004), and CCI (p = 0.042) (Table 2).

**Figure 1.**
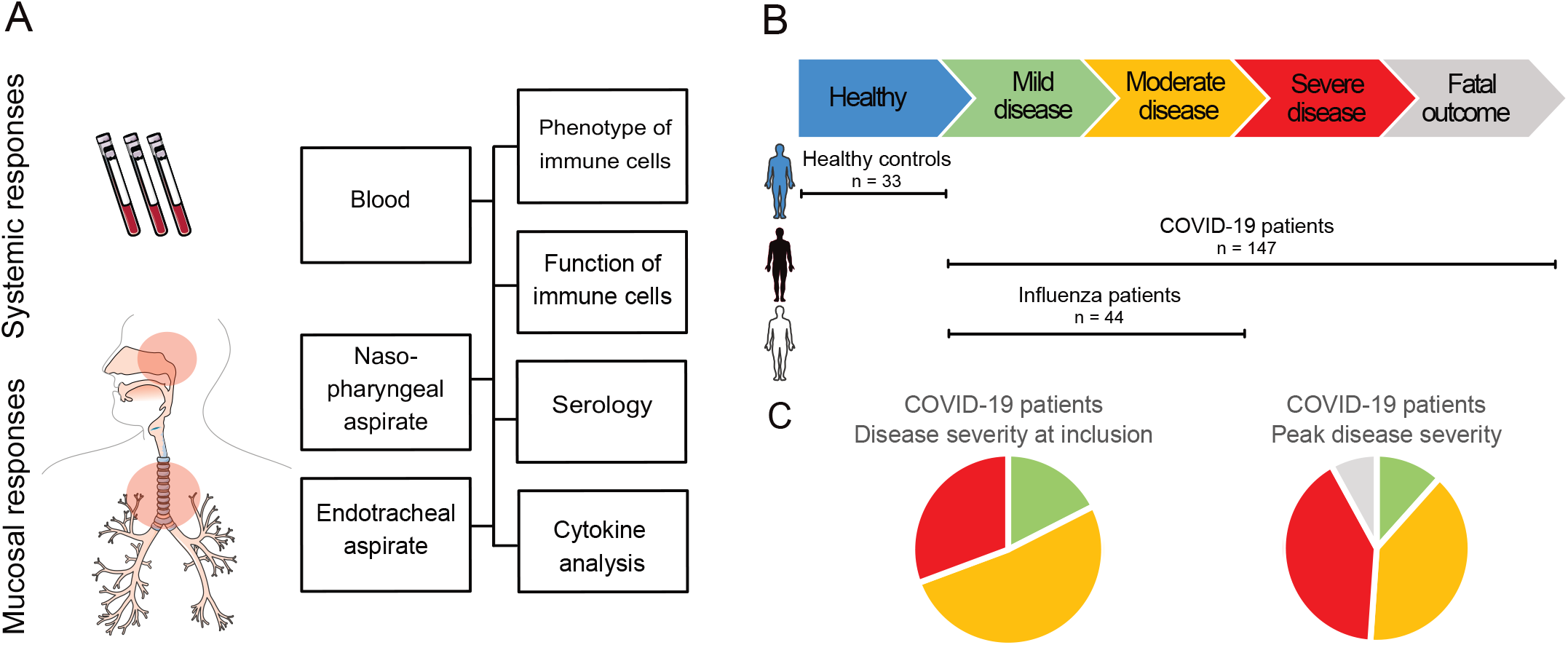
Study outline. (**A**) Blood and nasopharyngeal aspirates (NPA) were collected from COVID-19 patients, influenza patients and healthy controls. From ICU patients, endotracheal aspirates (ETA) were also collected. Cells were isolated from blood (PBMCs), NPA and ETA and were analyzed fresh using flow cytometry, and used for functional experiments. Aspirates and plasma were collected and used for serology and cytokine detection using ELISA. (**B**) Study subjects were included and sampled across disease severity, ranging from healthy controls (n = 33) to mild or moderate influenza disease (n = 44) and mild to fatal COVID-19 (n = 147). (**C**) Pie charts show distribution of disease severity at time of inclusion and the peak disease severity of COVID-19 patients. At time of inclusion: mild 18%, moderate 52% and severe 31%. At peak disease severity: mild 12%, moderate 39%, severe 41% and fatal outcome 8%.

**Table 1.** Patient and control characteristics

| **Cohort** | **COVID-19** | **Influenza** | **HC** | **Sig.<sup>1</sup>** |
| --- | --- | --- | --- | --- |
| n | 147 | 44 | 33 |  |
| Age, mean (range) | 57 (24-78) | 52 (18-88) | 51 (24-80) | * |
| Male gender, n (%) | 109 (74%) | 19 (43%) | 23 (70%) |  |
| Onset to admission, days, mean (SD) | 10 (5.8) | 5 (3.2) | - | * |
| Onset to inclusion, days, mean | 18 (10) | 5 (2) | - | *** |
| **Co-morbidities** |  |  |  |  |
| CCI, mean (SD) | 2 (2) | 2 (2) | - | ns |
| BMI, mean (SD) | 28.5 (4.6) | 26.7 (4.9) | 25.0 (3.3) | * |
| Hypertension, n (%) | 55 (38%) | 14 (33%) | 0 | ns |
| Diabetes, n (%) | 34 (23%) | 4 (9.1%) | 0 | ns |
| Current smoker, n (%) | 10 (7.0%) | 7 (16%) | 0 | ns |
| **Lab analyses** |  |  |  |  |
| CRP (mg/L), mean (SD) <sup>1,2</sup> | 206 (134) | 119 (146) | 2 (3) | *** |
| WBC (x10<sup>9</sup>/L), mean (SD) <sup>1,3</sup> | 8.0 (4.3) | 6.8 (3.5) | 6.5 (1.5) | ns |
| Lymphocytes (x10<sup>9</sup>/L), mean (SD) <sup>1,3</sup> | 0.89 (0.69) | 1.06 (0.47) | 2.41 (1.24) | *** |
| Neutrophils (x10<sup>9</sup>/L), mean (SD) <sup>1,3</sup> | 6.4 (3.9) | 5.1 (3.5) | 3.1 (0.7) | ** |
| Monocytes (x10<sup>9</sup>/L), mean (SD) <sup>1,3</sup> | 0.44 (0.27) | 0.64 (0.35) | 0.54 (0.24) | ** |
| **Outcome** |  |  |  |  |
| Peak severity score<sup>4</sup>, mean (SD) | 4.49 (1.65) | 1.80 (0.82) | - | *** |
| Fatal outcome | 12 (8.1) | 0 | 0 |  |

<sup>1</sup> Statistical tests performed: One-way ANOVA; Fisher's exact test; Pearson chi-square test. <sup>2</sup> Peak value <sup>3</sup> Value at maximum lymphopenia. <sup>4</sup> Peak severity score on 7-grade composite scale (see Methods).
Abbreviations: BMI = body mass index, CCI = Charlson co-morbidity index, CRP = C-reactive protein, WBC = White blood count.
Normal range: CRP <3 mg/L, WBC 3.5-8.8x10<sup>9</sup>/L, lymphocytes 1.1-3.5 x10<sup>9</sup>/L, neutrophils 1.6-5.9 x10<sup>9</sup>/L, monocytes 0.2-0.8 x10<sup>9</sup>/L.

**Table 2.**
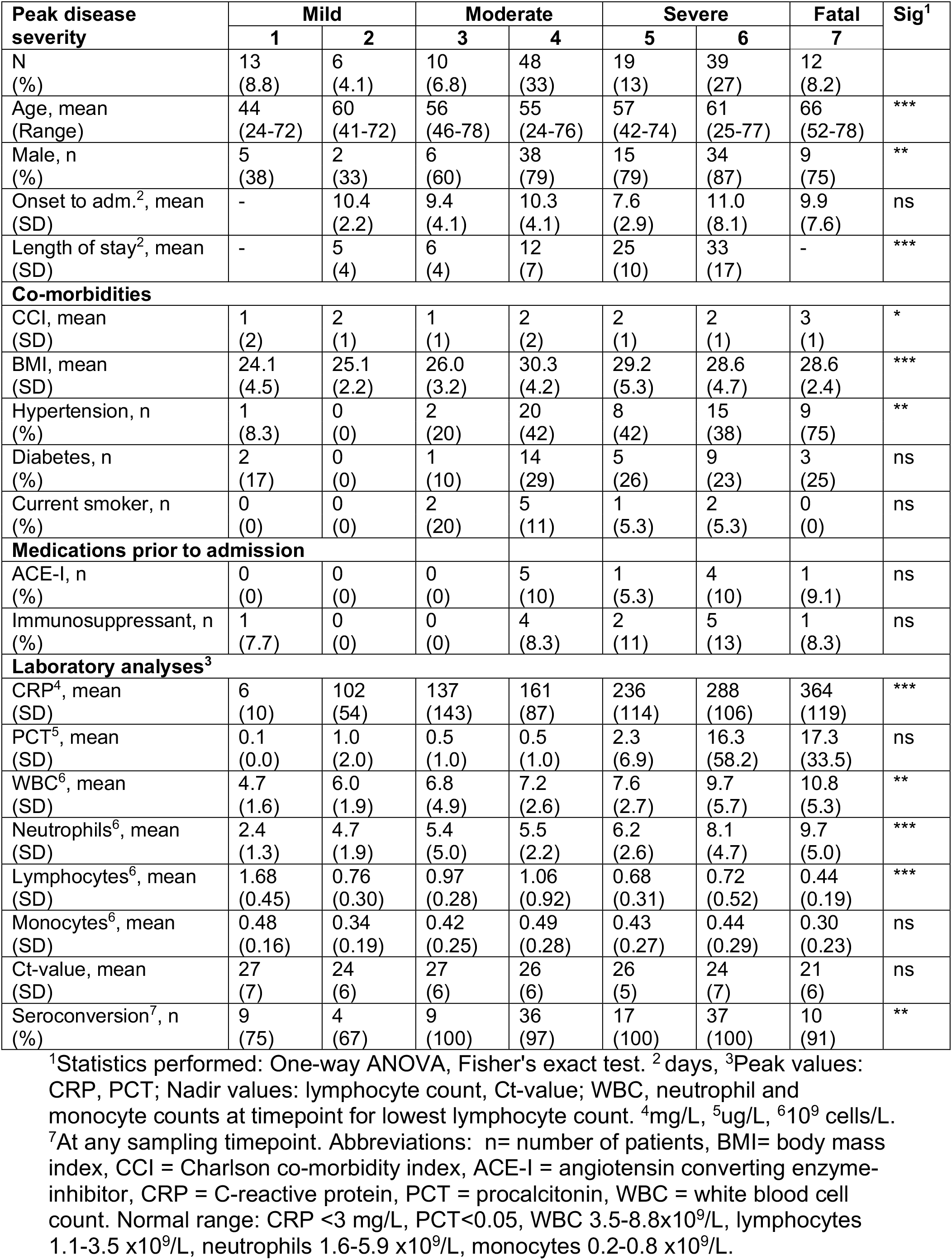
Baseline characteristics of COVID-19 patients across disease severity.

### M-MDSC frequencies are elevated in blood from COVID-19 and influenza patients proportional to disease severity

To investigate the dynamics of M-MDSCs during COVID-19 disease, we performed an extensive analysis of samples from COVID-19 patients across disease severity and compared with samples from influenza patients and HCs. PBMCs and cells from nasopharyngeal aspirates (NPA) and endothracheal aspirates (ETA) were stained and analysed by flow cytometry. M-MDSCs were identified as CD14+ cells within the lineage negative (CD3–CD56–CD19–CD20–CD66–), HLA-DR negative population (Figure 2A). In blood, the peak frequency of M-MDSCs was significantly increased in both COVID-19 patients and influenza patients compared to HCs (Figure 2B). The frequency of M-MDSC in NPA had a higher spread among both HC and patients compared to blood (Figure 2B). Albeit in a small number of patients with mild to moderate disease, influenza patients displayed a clear pattern of elevated frequencies of M-MDSCs in NPA as compared to COVID-19 patients and HCs. The elevated frequency of M-MDSCs in NPA in influenza patients compared to COVID-19 was also evident when comparing only mild and moderate influenza and COVID-19 cases, p = 0.0016 (data now shown). In contrast to NPA, COVID-19 patients with more severe disease had significantly higher peak M-MDSC frequencies in blood, while COVID-19 patients with mild disease had blood M-MDSC frequencies comparable to those seen in HCs (Figure 2C). The frequency of M-MDSC in blood from COVID-19 patients decreased over time (Figure 2D) and returned to similar frequencies as seen in HCs at follow up samples taken during convalescence (33–65 days after study inclusion) (Figure 2E).

**Figure 2.**
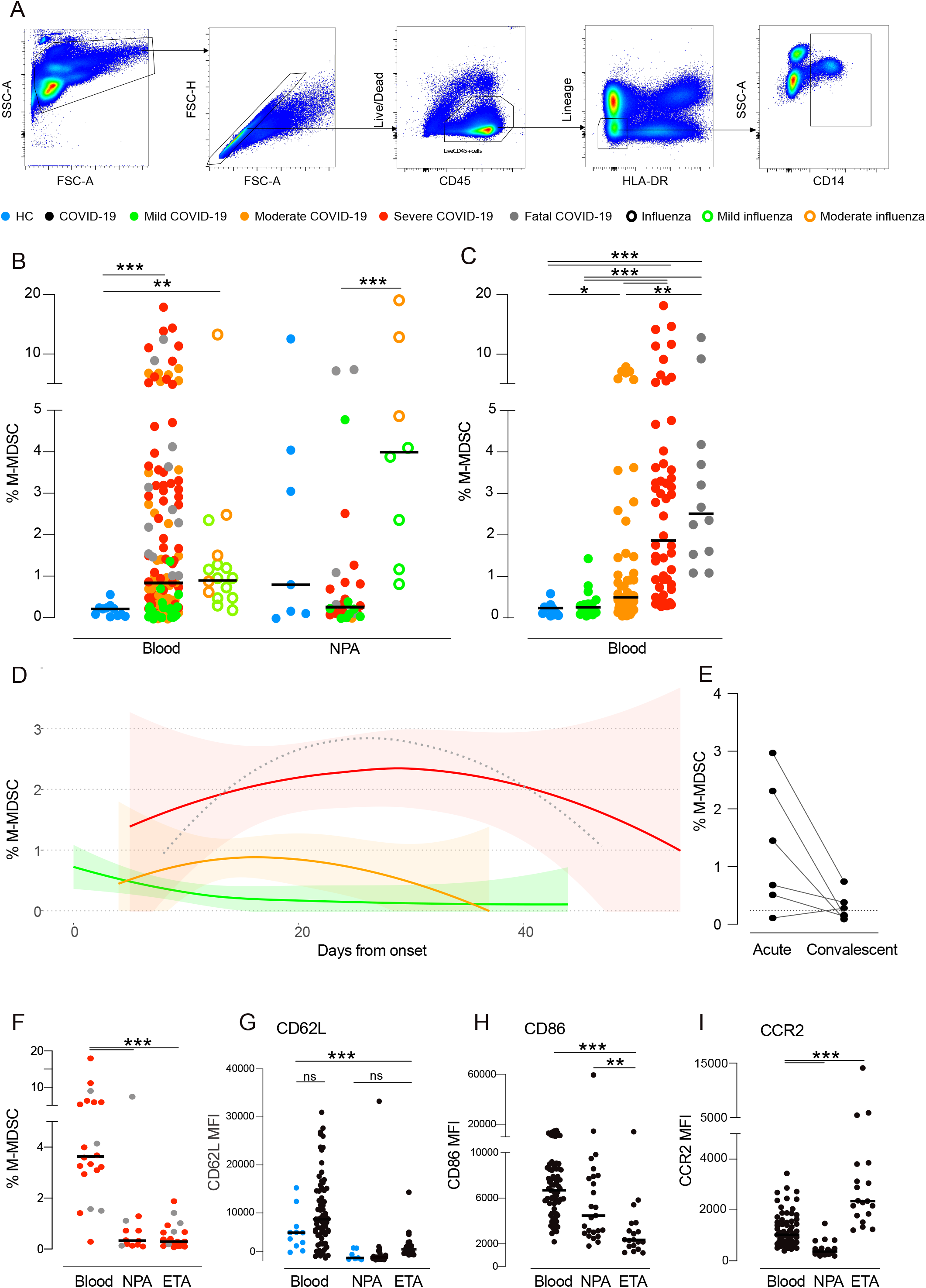
Frequency of respiratory and blood M-MDSC in COVID-19 patients, influenza patients and HCs. (**A**) Gating strategy to identify monocytic myeloiderived suppressor cells (M-MDSCs) by flow cytometry. Live, single CD45+ ukocytes were identified and cells expressing CD3, CD19, CD20, CD56, and CD66abce (lineage) and HLA-DR excluded. Among lineage-, HLA-DR- cells, CD14+ M-MDSCs were identified. (**B**) Frequency of M-MDSCs per live CD45+ cells in blood and NPA. HCs (blue): n = 12 (blood), 7 (NPA). Influenza patients (open circles): n = 19 (blood), 9 (NPA). COVID-19 patients (closed circles): n = 140 (blood), 28 (NPA). Points color-coded by peak severity. (**C**) Peak frequency of blood M-MDSCs per live CD45^+^cells across disease severity. HCs (blue): n = 12. COVID-19 patients (color-coded by peak severity): mild (n = 19), moderate (n = 53), severe (n = 56) and fatal (n = 12). **(D)** Blood M-MDSC frequency grouped by peak severity over time in COVID-19 patients. Line shows locally estimated scatterplot smoothing (LOESS) with shaded 95% CI (fatal group CI very wide, not presented) (**E)** Frequency of blood M-MDSCs in paired acute and convalescent samples from COVID-19 patients (n = 6). (**F**) M-MDSC frequency in blood, NPA and ETA samples from severe (red) and fatal (grey) COVID-19 patients. **(G-I)** Surface expression on M-MDSCs in HCs (blue) and COVID-19 patients (black) from blood, NPA and ETA (**G**) CD62L (**H**) CD86 (**I**) CCR2. **(B, C, F-I)** Comparisons of M-MDSC frequencies were performed using the non-parametric Kruskal-Wallis test and subsequent Dunn’s post-hoc test of multiple comparisons. In strip charts, group medians are presented as horizontal lines and individual patients as jitter points.

Somewhat surprisingly, COVID-19 patients, in contrast to influenza patients, had low frequencies of M-MDSCs in NPA, even when comparing patients with similar severity of disease (Figure 2B). Since COVID-19 patients on average were included in the study and sampled significantly later after onset of symptoms compared to the influenza patients (18 vs 5 days, respectively), we speculated that the infection-induced inflammation, including infiltration of M-MDSCs, might have transitioned into the lower airways in the COVID-19 patients compared to the influenza patients. To address this, we assessed whether M-MDSCs were present in ETA from the lower airways in 20 intubated COVID-19 patients. However, the frequency of M-MDSCs in ETA was not found to be elevated compared to NPA from the same patients (Figure 2F). Although the levels of blood M-MDSCs in COVID-19 patients were significantly elevated compared to HCs, the M-MDSC phenotype with respect to expression levels of CD62L, CCR2 and CD86 were similar (Figure 2G and data not shown). However, respiratory M-MDSCs expressed significantly lower levels of CD62L, CD86 and CCR2 compared to blood M-MDSCs (Figure 2G-I). Altogether, these data indicate that severe COVID-19 disease is associated with elevated levels of M-MDSCs in the blood, but not in the respiratory tract, at least at the time points studied.

### M-MDSCs isolated from COVID-19 patients suppress CD4 and CD8 T cell proliferation

To functionally confirm the identity of M-MDSCs in COVID-19 patients, we evaluated their suppressive effect on T cells. COVID-19 patient blood M-MDSCs were purified and co-cultured with CFSE-labelled allogeneic PBMCs, in the presence of SEB for three days (Supplementary figure 3). As expected, SEB induced strong T cell proliferation (Figure 3A). However, addition of M-MDSCs induced a significant suppression of both CD4 and CD8 T cell proliferation in a dose-dependent manner (Figure 3A-C). In line with this, SEB induced high levels of IFNγ secretion that were significantly lower in co-cultures with M-MDSCs present (Figure 3D). Arginase 1 (Arg-1) production is one effector mechanism by which M-MDSCs suppress T cell proliferation via degradation of L-arginine that is needed for proliferation. Indeed, addition of L-arginine to the M-MDSC co-cultures restored the concentration of IFNg in the cell culture supernatants (Figure 3D). Furthermore, co-cultures with M-MDSCs contained high levels of Arg-1 that was undetectable in cultures without M-MDSCs (Figure 3E). In co-cultures supplemented with L-arginine, Arg-1 was no longer detectable, possibly due to complex formation of Arg-1 and the substrate^25^ (Figure 3E). Importantly, the addition of recombinant L-arginine to the co-cultures decreased the suppressive effect of M-MDSCs on T cells and partially restored T cell proliferation (Figure 3F-G). This indicates that M-MDSCs from COVID-19 patients use Arg-1 as one mechanism to suppress T cells. In conclusion, blood M-MDSC isolated from COVID-19 patients are functional and can suppress T proliferation and IFNγ secretion in a dose- and Arg-1 dependent manner.

**Figure 3.**
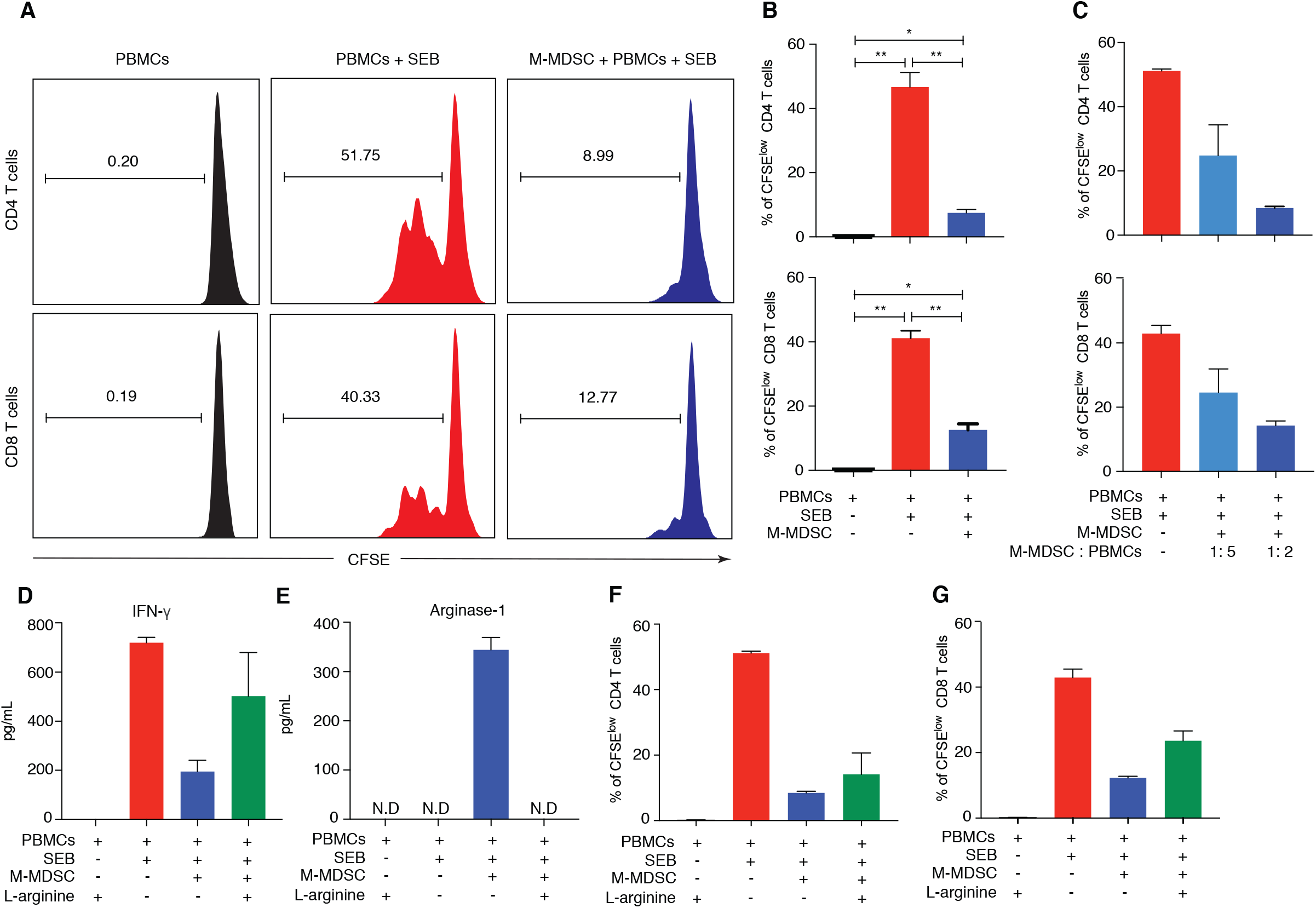
M-MDSCs isolated from COVID-19-patients suppress T cell proliferation partly through release of Arginase-1. (**A**) Blood M-MDSC isolated from COVID-19 patients were co-cultured with CFSE-labeled allogenic PBMCs in the presence of SEB for 3 days, at a ratio of 1:2 (M-MDSC:PBMC). Histograms show representative CD4 and CD8 T cell proliferation as assessed by CFSE dilution and flow cytometry. Number indicate frequency of proliferating T cells. (**B**) Bar graphs showing percentage of proliferating CD4 and CD8 cells with mean ± SEM (n = 3). Statistical testing performed using Wilcoxon signed ranks test. (**C**) Isolated M-MDSC were cultured with CFSE-labelled allogenic PBMCs in the resence of SEB for 3 days. The ratio of M-MDSC: PBMC were 1:5 and 1:2. Bar graphs show percentage of proliferating CD4 T and CD8 cells with mean ± SEM (n = 2). (**D**) Bar graphs show levels of IFNg in supernatants from cell cultures with mean ± SEM (n = 2). (**E** to **G**) Isolated M-MDSC were cultured with CFSE-labelled allogenic PBMCs in the presence of SEB and L-arginine for 3 days. The ratio of M-MDSC: PBMC was 1:2. (**E**) Bar graphs show levels of Arginase-1 in supernatants from cell cultures with mean ± SEM (n = 2). (**F** and **G**) Bar graphs show percentage proliferating (**F**) CD4 T cells and (**G**) CD8 T cells (n = 2) with mean ± SEM (n = 2).

### M-MDSC related cytokines are elevated in COVID-19 patients and increase with disease severity

To further investigate the effect of elevated frequencies of blood M-MDSCs in COVID-19 patients, cytokines that have been linked to M-MDSC function and activation were measured in plasma and NPA at the time of study inclusion. In plasma, COVID-19 patients had significantly higher levels of Arg-1 than HCs, but no significant difference was observed between COVID-19 patients and influenza patients (Figure 4A). Interestingly, Arg-1 levels in NPA were higher than in plasma in all three groups, with no significant differences between the groups (Figure 4B). Among COVID-19 patients, the plasma concentration of Arg-1 was lower in patients with mild disease compared to patients with moderate, severe and fatal disease (p = 0.07, p = 0.04 and p = 0.01 respectively) (Figure 4C).

**Figure 4.**
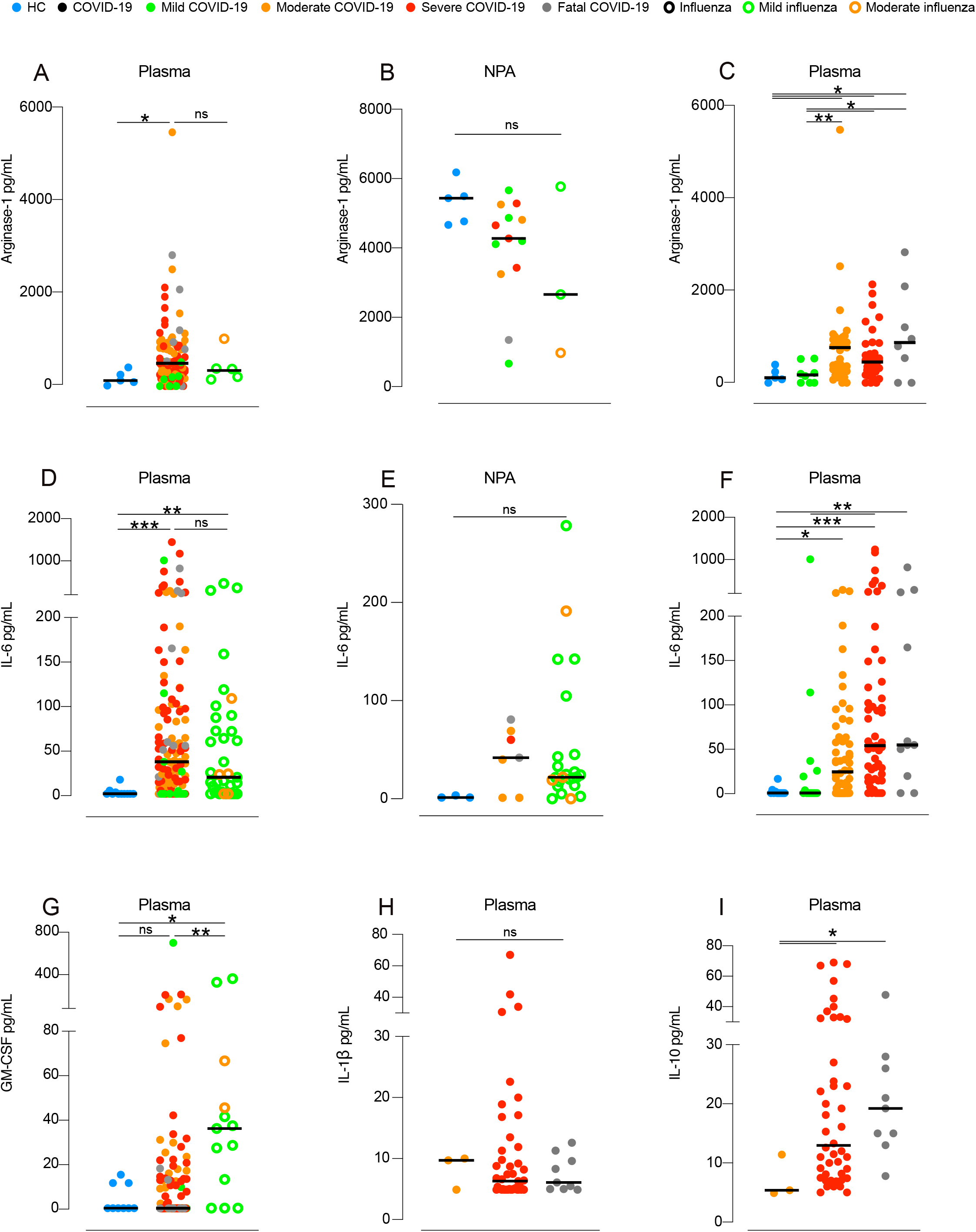
Levels of cytokines in blood and NPA from HCs, COVID-19 patients and influenza patients. **(A-C)** Arginase-1 was measured in **(A)** plasma, (**B**) NPA and (**C**) across COVID-19 disease severity in plasma. HCs (blue points): n = 5 (blood), 5 (NPA). Influenza patients (open circles): n = 6 (blood), 3 (NPA). COVID-patients (closed circles): n = 93 (blood), 13 (NPA). COVID-19 patients (colorcoded by peak severity): mild (n = 8), moderate (n = 41), severe (n = 36) and fatal (n = 8). **(D-F)** display IL-6 as measured in **(D)** plasma, (**E**) NPA and (**F**) IL-6 across COVID-19 disease severity in plasma. HCs (blue points): n = 11 (blood), 3 (NPA). Influenza patients (open circles): n = 37 (blood), 24 (NPA). COVID-19 patients (closed circles): n = 133 (blood), 7 (NPA). COVID-19 patients (color-coded by peak severity): mild (n = 14), moderate (n = 56), severe (n = 52) and fatal (n = 11). (**G**) GMCSF in plasma comparing HCs and patients. **(H**) IL-10 and **(I**) IL-1B in plasma from patients with moderate to severe disease or fatal outcome. HCs (blue points): n = 9. Influenza patients (open circles): n = 13. COVID-19 patients (closed circles): n = 106. COVID-19 patients (color-coded by peak severity): mild (n = 12), moderate (n = 38), severe (n = 48) and fatal (n = 8). **(A-I)** Medians were compared using the nonparametric Kruskal-Wallis test. Post-hoc testing was carried out while controlling the False Discovery Rate **(A-C)** or by using Dunn’s test of multiple comparisons (**D-I**). In strip charts, group medians are presented as horizontal lines and dividual patients as jitter points.

Plasma concentrations of IL-6, a potent proinflammatory cytokine important for M-MDSC differentiation^14^, were significantly increased in both COVID-19 patients and influenza patients compared to HCs (Figure 4D), while no statistically significant differences were observed in NPA (Figure 4E). Furthermore, levels of IL-6 were strikingly different across the COVID-19 disease severity groups (Figure 4F). Mild COVID-19 patients had significantly lower IL-6 levels than moderate patients (p = 0.038), severe patients (p< 0.001) and patients with fatal outcome (p = < 0.01), respectively. COVID-19 patients with moderate disease also had significantly lower levels than severe patients (p = 0.04). GM-CSF, which is important for M-MDSC development^14^, was also measured in plasma, but was only significantly elevated in influenza patients (Figure 4G). Finally, concentrations of IL-10 and IL-1β were measured in plasma of a subset of COVID-19 patients with moderate to fatal disease, but only IL-10 showed an association with disease severity (Figure 4H-I). In summary, cytokines involved in the activation and function of M-MDSCs were elevated in plasma from COVID-19 patients, and correlated with disease severity.

### T cells are reduced in blood of COVID-19 patients and have low CD3ζ chain expression

Since M-MDSCs isolated from COVID-19 patients efficiently suppressed T cells *in vitro*, the overall blood T cell frequency and function in patients was assessed in the same patients using flow cytometry (Supplementary figure 4). Absolute numbers of peripheral blood CD4+ T cells were decreased in the COVID-19 patients with moderate, severe and fatal disease compared to HCs (p = 0.02, p = 0.001, p = 0.05 respectively) (Figure 5A). Similarly, absolute numbers of CD8+ T cells were also significantly decreased in moderate, severe and fatal COVID-19 disease compared to HCs (p = 0.004, p = 0.005 and p = 0.003 respectively) (Figure 5B). However, no correlation between blood M-MDSC frequency and T cell count in COVID-19 patients was found (Figure 5C), either at peak/bottom frequencies or at any of the longitudinal time points studied in each patient (Figure 5C and data not shown).

**Figure 5.**
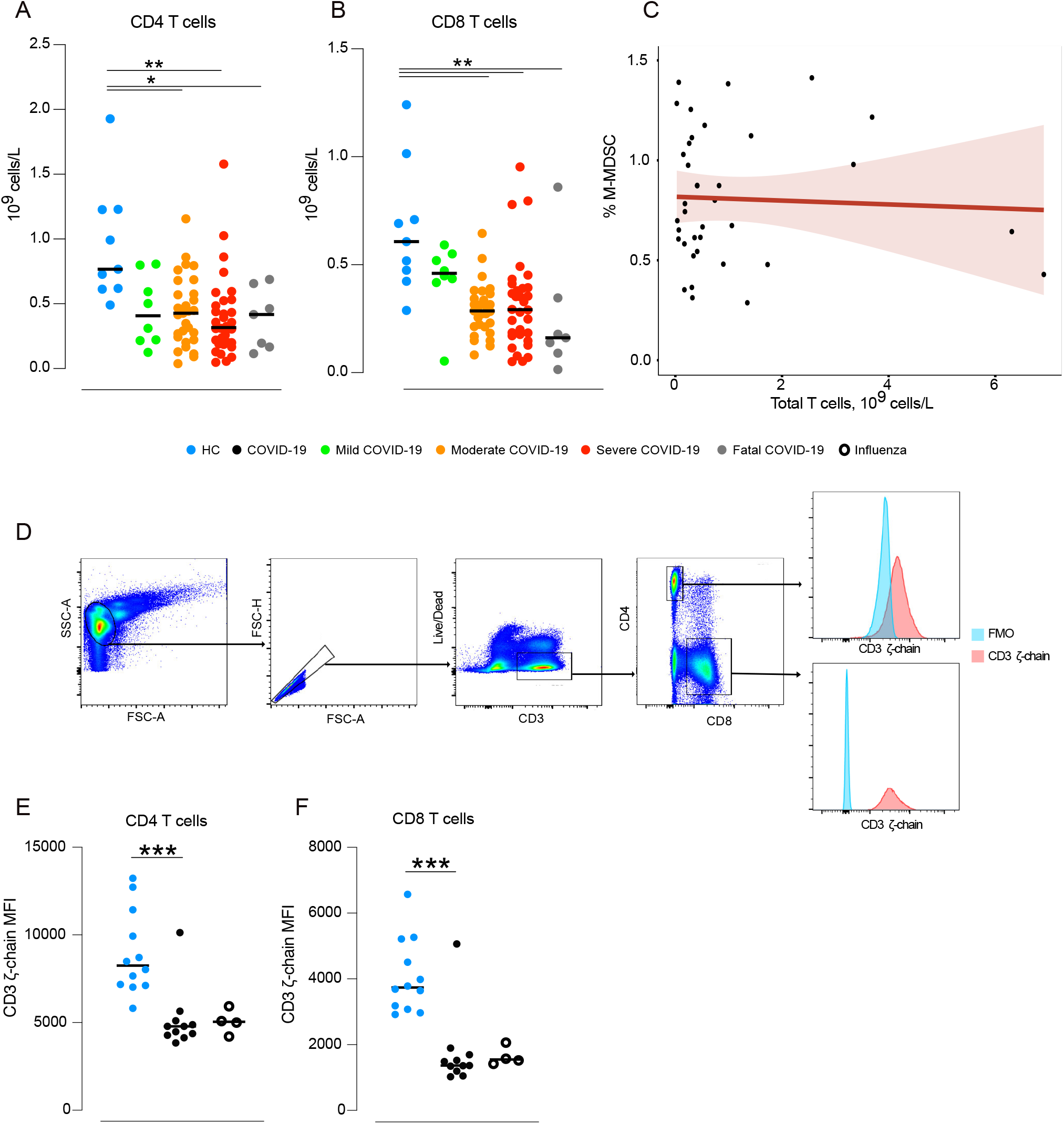
T cells in COVID-19 patients. **(A-B)** Total lowest CD4 and CD8 T cell count in blood was calculated in HCs (n = 9) and COVID-19 patients across disease severity, mild (n = 8), moderate (n = 29), severe (n = 32), fatal (n = 7). **(C)** Spearman correlation between total CD3+ T cell count in blood and peak M-MDSC frequency. **(D)** Gating strategy for CD3ζ chain analysis. Live, single CD3+ cells were identified, and separated into CD4+ or CD8+ T cells. MFI was calculated for both populations. The blue histogram represents the fluorescence-minus-one control (FMO) and the red histogram represents CD3ζ (PE-labelled). (**E-F**) Thawed PBMCs from 11 COVID-19 patients, 4 influenza patients and 12 matched healthy controls were intracellularly stained for CD3ζ chain expression. Median fluorescent intensity (MFI) was calculated for (**E**) CD4+ and (**F**) CD8+ T cells. Comparison of medians between groups was performed using Kruskal-Wallis’ test and subsequent post-hoc testing by Dunn’s test of multiple comparisons.

Evidence of T cell suppression was further investigated in COVID-19 patients with varying disease severity, influenza patients and HCs by quantifying the surface expression of the CD3ζ chain, a homodimer chain in the TCR complex involved in T cell proliferation and in secretion of cytokines (Figure 5D). The CD3ζ chain is downregulated *in vitro* in the absence of L-arginine resulting in decreased T cell proliferation^26^. We observed that the surface expression of the CD3ζ chain on CD4+ and CD8+ T cells was significantly lower in both COVID-19 and influenza patients compared to HC (Figure 5E-F) suggesting that the T cells may have an impaired functional capacity. In summary, COVID-19 patients had lower T cell frequency and indication of impaired function of the T cells compared to HCs.

### Early M-MDSC frequencies predict peak disease severity

To evaluate the effect of blood M-MDSC frequency early during COVID-19 disease on subsequent disease severity, a proportional odds logistic regression was performed (Figure 6A). This yielded a crude odds ratio (OR) of 1.50 (95% CI 1.03 – 2.32), indicating that the M-MDSC frequency in the first two weeks from onset of symptoms could potentially be a predictor of more severe outcome of COVID-19 (Figure 6B).

**Figure 6.**
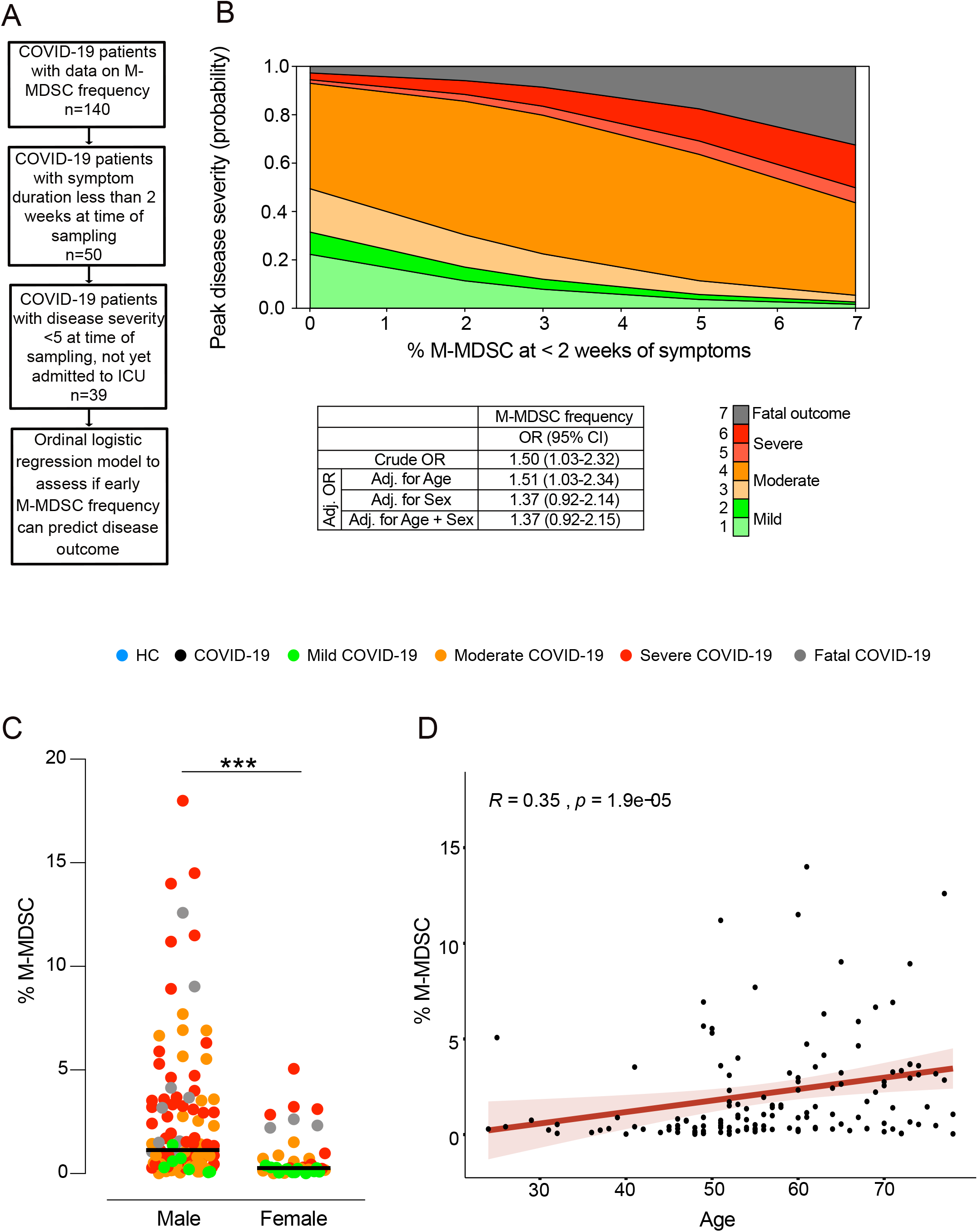
M-MDSC frequency predicts disease severity and is associated with male gender and age. **(A)** Criteria for inclusion in the ordinal logistic regression model. Only patients with up to 2 weeks of symptoms and disease severity < 5 that had not yet been admitted to the ICU were included in the analysis **B**) Proportional odds logistic regression showing predictive capacity of M-MDSC-frequency on peak disease severity score. Crude and adjusted odds ratio presented. (**C**) Peak M-MDSC frequency in men and women compared using Wilcoxon-Mann-Whitney-U (**D**) Spearman correlation between age and peak M-MDSC frequency.

As shown above, M-MDSC frequency was higher in COVID-19 patients with more severe disease (Figure 2C), and these patients were both predominantly male and had a significantly higher age compared to COVID-19 patients with less severe disease (Table 1). Therefore, the association between M-MDSC frequency and sex and age was assessed, demonstrating significantly higher levels of M-MDSCs in men (Figure 6C) and a significant correlation between age and M-MDSC frequency (R = 0.22, p = 0.01) (Figure 6D).

In summary, early M-MDSC frequencies are associated with subsequent disease severity and appear to be strongly associated with age and sex.

## Discussion

Understanding the immunopathogenesis of COVID-19 is critical to optimally treat patients and prevent fatal outcome but also to aid in the development of specific therapy and vaccines. One potential player in the immune activation to SARS-CoV-2 infection may be MDSCs, a subset of immune cells that in recent years have been intensively studied, by us and others, mainly in relation to cancer and vaccination^14, 15, 16, 18, 27^. Still, only limited knowledge on how M-MDSCs influence disease severity during infection, including COVID-19, is available. In the present study, the distribution and function of M-MDSCs during COVID-19 were investigated over time and across disease severity in a comparatively large and clinically well-characterized cohort.

A major finding in the current study is the association between M-MDSC frequency and disease severity in COVID-19 patients. Downregulation of HLA-DR on monocytes has previously been shown in severe COVID-19, possibly reflecting an increase in M-MDSC frequency, and is linked to high levels of IL-6 and lymphopenia^28^. We observed an increased frequency of blood M-MDSC, both during COVID-19 and influenza, similar to what was previously observed in HIV-1^17, 29^. While the COVID-19 and the influenza patient cohorts are not completely comparable in disease severity and time of sampling after symptom onset, it is still relevant to know that expansion of M-MDSCs is also observed in another acute respiratory infection, and that this finding is not unique for COVID-19. Furthermore, MDSCs are expanded during sepsis, and are associated with poor outcome^30^.

Notably, in influenza patients M-MDSC frequencies were increased in the nasopharynx compared to blood, indicating that M-MDSCs are recruited to the site of infection during influenza similar to what has been observed for other myeloid cells^31^. In contrast, the frequency of M-MDSC in the nasopharynx of COVID-19 patients was similar to HCs. Although SARS-CoV-2 replication is initiated in the upper airways, it frequently progresses to the lower respiratory tract^32, 33^, and may result in immune cell recruitment in the lower airways. However, we were unable to find higher frequencies of M-MDSCs in ETA than in NPA. Nevertheless, ETA does not necessarily reflect cells in the alveoli, but is rather a reflection of the trachea and bronchi. It is therefore possible that M-MDSCs are present even deeper down in the lungs and/or that the time points we sampled the COVID-19 patients were past the peak accumulation of M-MDSCs in the airways. Alternatively, M-MDSCs could differentiate to more macrophage-like cells at the site of inflammation, as seen after migration to tumor sites^34, 35^, and not be identified with the flow cytometry staining panel used. Migration of M-MDSC from blood to the site of infection is supported by the fact that M-MDSCs in blood had upregulated CD62L compared to respiratory M-MDSCs. CD62L, or L-selectin is involved in the extravasation of immune cells^36^. Continued studies are ongoing to further address the kinetics and mechanism of myeloid cell migration in humans.

Characterizing MDSCs is challenging due to the lack of unique cell surface markers and therefore functional analysis of the cells is critical to validate phenotypic identification^15, 19^. We verified the suppressive capacity of the M-MDSCs in COVID-19 patients and found that HLA-DR- CD14+ cells isolated from COVID-19 patients had a potent suppressive effect on T cells, demonstrating that the M-MDSCs identified by flow cytometry corresponded to suppressive and functionally active cells. Since Arg-1 was important for M-MDSC activity *in vitro*, levels were measured in plasma and NPA from patients. As expected, COVID-19 patients had increased level of Arg-1 compared to HCs in plasma. There was also a connection between disease severity and Arg-1 level in plasma, although there was no difference between patients with moderate disease to fatal outcome. Interestingly, Arg-1 levels were in general substantially higher in NPA than in plasma for all cohorts, without any association with M-MDSC frequency in NPA. It is known that Arg-1 is constitutively produced in the airways in bronchial epithelial cells, endothelial cells, myofibroblasts and alveolar macrophages. The function, however, is unknown. Arg-1 has been suggested to be involved in regulation of NO and airway responsiveness and tissue repair^25^. The association between M-MDSC and Arg-1 in the airways merits further examination.

Several factors are involved in the expansion of M-MDSCs, including IL-6 and GM-CSF^14^. In line with previous studies^12, 28^, we demonstrated a relationship between IL-6 and COVID-19 disease severity. The high levels of IL-6 could contribute to the generation of M-MDSCs, especially in patients with severe disease. In contrast to previous reports^37^, plasma GM-CSF was not elevated in our COVID-19 cohort. Instead, higher levels of GM-CSF were seen in the influenza patient cohort. This could be explained by kinetics: the COVID-19 group had longer duration of symptoms compared to the influenza patients and it is possible that the level of GM-CSF had already decreased. IL-10 is also relevant in respect to MDSCs, since it can be produced by MDSCs^14, 18^, and plasma levels increased with rising disease severity.

A decrease in absolute numbers of both CD4+ and CD8+ T cells, in line with the data presented here, has previously been demonstrated in COVID-19 patients with severe disease, but the underlying mechanisms for the decrease are still unknown^38^. One potential such mechanism could be the downregulation of CD3ζ chain, that results in impaired T cell proliferation. CD3ζ chain downregulation on T cells has previously been observed in relation to MDSCs in several conditions including sepsis, hepatitis C infection and gastric cancer^39, 40, 41^. By establishing an immunosuppressive environment, M-MDSCs might prevent efficient immune activation and impede the development of specific adaptive responses required to clear the infection. We therefore speculate that expansion of M-MDSCs contributes to the immune imbalance described in COVID-19, possibly favouring disease progression.

There is an urgent need for a better understanding of the pathophysiology of COVID-19, including predictors of disease severity. In the current study, we investigated whether M-MDSC frequencies in blood early in disease (before patients develop severe disease) could affect disease outcome. Although our analysis should be interpreted with caution because of low power and possible risk of overfitting when adjusting for age and sex, it clearly suggests elevated M-MDSC frequency in the first two weeks from onset of symptoms as a risk factor for poorer disease outcome. Our protein and functional M-MDSC COVID-19 data is in line with emerging RNA seq data that identify a population likely to be M-MDSCs that is associated with severe COVID-19^21^. Previous research has shown age and male gender are risk factors for severe COVID-19^42^. Furthermore, it is known that MDSCs increase with age as part of the “inflammaging” process (the inflammation seen in ageing)^43^, but little is known about gender differences. In line with this, we found a correlation between M-MDSC frequency and both age and male gender in this cohort, possibly partly explaining the increased morbidity seen in men and older patients.

In summary, we have shown that M-MDSCs are expanded in both COVID-19 and influenza and are associated with disease severity in COVID-19 patients, especially in men and older individuals. Although we found indirect indications of M-MDSC migration, we were unable to show an increased M-MDSC frequency in respiratory mucosa in the same way as in influenza, presenting an area of future research. In this study we also demonstrated that M-MDSCs isolated from COVID-19 patients are effective T cell suppressors *in vitro* and may partly explain the detrimental decrease of T cells in patients with severe COVID-19. Finally, we showed that M-MDSCs may predict disease outcome. The findings of this study provide yet another important piece to the puzzle of fully comprehending the immunologic profile of COVID-19 and can potentially contribute to future therapeutic and diagnostic advancements.

## Methods

### Ethics statement

The study was approved by the Swedish Ethical Review Authority, and performed according to the Declaration of Helsinki. Written informed consent was obtained from all patients and controls. For sedated patients, the denoted primary contact was contacted and asked about the presumed will of the patient and, if applicable, to give initial oral and subsequently signed written consent. When applicable retrospective written consent was obtained from patients with non-fatal outcomes.

### Study subjects

Inclusion of COVID-19 patients was performed at Karolinska University Hospital and Haga Outpatient Clinic (Haga Närakut), Stockholm, Sweden during March-May 2020. Inclusion was performed at various levels of care, ranging from primary to intensive care. Inclusion criteria were age > 18 years and PCR-confirmed SARS-CoV-2 infection. In order to recruit mild/asymptomatic cases, household contacts of COVID-19 patients were screened with PCR and recruited if positive. Similarly, adult patients with PCR-confirmed influenza A virus infection were recruited during January-March 2019 and 2020. A cohort of healthy controls (HCs) (i.e. confirmed influenza A virus and SARS-CoV-2 negative by PCR) was recruited and sampled in the same way as study patients.

Degree of respiratory failure was categorized daily according to the respiratory domain of the Sequential Organ Failure Assessment score (SOFA)^44^. If arterial partial pressure of oxygen (PaO_2_) was not available, peripheral transcutaneous hemoglobin saturation (SpO_2_) was used instead and the modified SOFA score (mSOFA) was calculated^45^. Fraction of inspired oxygen (FiO_2_) estimation based on O_2_ flow was done in accordance with the Swedish Intensive Care register definition as defined at (https://icuregswe.org/globalassets/riktlinjer/sofa.pdf, accessed on 7 Sep. 20). Patients were subsequently categorized based on the peak respiratory SOFA or mSOFA value. The 5-point respiratory SOFA score was then extended with an additional level to distinguish admitted mild cases from non-admitted mild cases. Finally, fatal outcome was added as a seventh level, with peak disease severity score 6 prior to death in all but two patients who had scores of 4 and 5, respectively. Additionally, the resulting 7-point composite peak severity score was condensed into a classification consisting of mild (1-2), moderate (3-4), severe (5-6), and fatal (7) disease (Supplementary Tables 1 and 2).

Medical records were reviewed for clinical history, laboratory analyses, medications, previous diseases and co-morbidities, and risk factors. Total burden of comorbidities was assessed using the Charlson co-morbidity index (CCI)^46^.

### Collection of respiratory and blood samples

Nasopharyngeal aspirates (NPA) were collected from COVID-19 and influenza patients and HCs where possible, and endotracheal aspirates (ETA) were collected from intubated COVID-19 patients in the ICU. Venous blood was collected in EDTA-containing tubes from all non-ICU patients and controls. In ICU patients, blood was pooled from heparin-coated blood gas syringes discarded in the last 24 hours. In some ICU patients, additional venous blood samples were also collected in EDTA tubes. Routine clinical chemistry analysis was performed on all study subjects including HCs. Admitted patients were sampled at up to four timepoints and discarded ICU patient material was collected up to ten timepoints.

### Isolation of cells from blood and nasopharyngeal aspirates

NPA samples were centrifuged at 400g/5 min/ room temperature (RT). Supernatant was collected and frozen at –20°C. Cells were washed with sterile PBS and mucus was removed using a 70 µm cell strainer. Blood samples were centrifuged at 800g/8 min/ RT. Plasma was collected and frozen at –20°C. The cellular fraction was diluted with sterile PBS and PBMCs isolated by density-gradient centrifugation at 900g/25 min/RT (without brake), using Ficoll-Paque Plus (GE Healthcare). Cell count and viability were assessed using Trypan Blue (Sigma) exclusion with an automated Countess cell counter (Invitrogen). Cells were stained fresh for flow cytometry analysis. Excess PBMCs were cryopreserved in FBS (Gibco) with 10% DMSO (Sigma) and stored in liquid nitrogen.

### Flow cytometry

Cells were stained using Live/Dead Blue (Invitrogen), incubated with human FcR blocking reagent (Miltenyi Biotec) and stained with antibodies against the following surface proteins: CD1c (AD5-8E7; Miltenyi Biotec), CD3 (SK7; BD), CD11c (B-Ly6; BD), CD14 (M5E2; BD), CD16 (3GE; BioLegend), CD19 (HIB19; BioLegend), CD20 (L27; BD), CD45 (HI30; BD), CD56 (HCD56; BD), CD62L (SK11; BD), CD66abce (TET2; Miltenyi Biotec), CD86 (2331; BD), CD123 (7G3; BD), CD141 (AD5-14H12; Miltenyi Biotec), CCR2 (K036C2; BioLegend), CCR7 (150503; BD) and HLA-DR (TU36; Life Technologies). If enough cells were available, a second staining was performed using CD3 (SP34-2; BD), CD4 (L200; BD), CD11c (B-ly6; BD), CD14 (M5E2; BD), CD16 (3G8; BD), CD19 (SJ25-C1; Thermo Fisher Scientific), CD45 (HI30; BD), CD56 (HCD56; BioLegend), CD66abce (TET2; Miltenyi Biotec), CD123 (7G3; BD), LOX-1 (15C4; BioLegend) and HLA-DR (L243; BioLegend). All stainings were performed at 4°C for 20 minutes. Cells were washed with PBS and fixed with 1-2% paraformaldehyde.

The expression of CD247 (TCRζ, CD3ζ) on CD4 and CD8 T cells was evaluated by intracellular staining. Briefly, following surface staining with Live/Dead Blue (Invitrogen), CD3 (SP34-2; BD), CD4 (L200; BD) and CD8 (SK1; BD), cells were fixed and permeabilized with permeabilization buffer (Thermo Fisher Scientific) and then stained with anti-CD247 (6B10.2; BioLegend) at 4°C for 20 min. Samples were acquired on LSRFortessa flow cytometer (BD Biosciences). Data were analysed using FlowJo software 10.5.3 (TreeStar). Absolute numbers of CD4 and CD8 T cells were calculated by multiplying the frequency of T cells out of total lymphocytes obtained from flow cytometry data with the lymphocyte count from differential cell counts. If multiple T cell frequencies were available from the same patient, the lowest T cell count was used. If absolute lymphocyte count was missing, a value was linearly interpolated between existing values if no more than 7 days apart.

### M-MDSC T cell suppression assay

M-MDSCs (HLA-DR^−^ CD14+ cells) were purified from frozen PBMCs of three COVID-19 patients, following a protocol developed by Lin et al^16^. HLA-DR+ cells were depleted using anti-HLA-DR microbeads and an LD column. From the negative fraction, CD14+ cells were positively selected using anti-CD14 microbeads. MS columns and MACS separators (all Miltenyi Biotec) were used for the cell sorting. Approximately 0.2 million M-MDSCs were obtained from 25-30 million PBMCs, with a viability > 90% and purity > 85%. In parallel, cryopreserved PBMCs from a buffy coat were thawed and 4 million cells were labelled with CFSE (Thermofisher). The previously purified M-MDSCs were co-cultured with 0.5 million of the CFSE-labelled PBMCs, at a ratio of 1:2 or 1:5, in the presence of 0.1 µg/mL staphylococcal enterotoxin B (SEB) (Sigma-Aldrich) or 200µg/mL L-arginine (Sigma-Aldrich). The cells were incubated for 3 days at 37°C, in RMPI 1640 (Sigma-Aldrich) medium supplemented with 10% FCS, 5 mM L-glutamine, 100 U/mL penicillin and streptomycin (all Invitrogen). Supernatants were collected from cultures to measure secreted arginase-1 (Invitrogen) and IFNg (R&D Systems) by ELISA. The cells were washed and surface-stained with CD3 (SK7), CD4 (OKT4), and CD8 (SK1) (all from BD Biosciences). Flow cytometry (LSRFortessa, BD Biosciences) was performed as described above and T cell proliferation was measured, by calculating the percentage of CFSE^low^ T cells.

### Cytokine analysis

Cytokine levels were measured in plasma samples, NPA supernatants and culture supernatants using enzyme-linked immunosorbent assay (ELISA). IL-6, GM-CSF and IFNg ELISAs were performed using DuoSet® kits (R&D Systems). Arginase-1 ELISA was performed using Arginase-1 Human ELISA Kit (ThermoFisher). Levels of IL-10 and IL-1b were analyzed at Karolinska University Laboratory using Roche Cobas e602.

### Serology

Antibodies against the SARS-Cov-2 Spike (S) trimer were assessed by ELISA. Recombinant proteins were received through the global health-vaccine accelerator platforms (GH-VAP) funded by the Bill and Melinda Gates foundation. Briefly, 96-well plates were coated with 100ng/well of S protein. Plates were incubated with a selected duplicate dilution (1:50) of each plasma sample at ambient temperature for 2 hours.

Detection was performed with a goat anti-human IgG HRP-conjugated secondary antibody (clone G18-145 from BD Biosciences) followed by incubation with TMB substrate (BioLegend; cat# 421101) and stopped with a 1M solution of H_2_SO_4_. Absorbance was read at 450nm+550nm background correction using an ELISA reader. Data are reported as the average optical density (OD) value of the two duplicates.

### Statistical analysis

Data analysis was performed in RStudio version 1.2 (RStudio Inc., Boston, MA), GraphPad Prism version 8.0 (GraphPad Software Inc., San Diego, CA) and Microsoft Excel (Microsoft Corp., Redmond, WA). Routine analyses excluding cytokines and flow-cytometry data were assumed to have a standard distribution and means were compared using either an independent Student’s t-test or a one-way analysis of variance (ANOVA). Nominal patient characteristics were compared between groups using a Pearson Chi-Squared test or Fisher’s exact test depending on if the expected count of any cell was above or below five. Cytokine and flow cytometry data were presumed to have a non-standard distribution and medians were thus compared using the Wilcoxon-Mann-Whitney U or Kruskal-Wallis tests depending on number of cohorts. Post-hoc testing after Kruskal-Wallis was performed using Dunn’s test of multiple comparisons or by controlling the False Discovery Rate using Benjamini, Krieger and Yekutieli’s an adaptive linear step-up procedure. Non-parametric comparisons of dependent data were performed using Wilcoxon’s Signed Ranks test. For correlation analyses of continuous data, Spearman’s Rho was used. Finally, a proportional odds logistic regression model was constructed to evaluate predictive capacity of M-MDSC frequencies on disease severity. A 95% significance level was used throughout the study.

Missing daily severity score data was approximated using a last-observation-carried-forward (LOCF) method. For flow cytometry data, the peak M-MDSC frequency and lowest T cell count were extracted. The peak value for routine laboratory analyses was extracted separately for each analysis except in the case of blood differential counts in which all counts were extracted for the timepoint of lowest lymphocyte count. The ordinal logistic regression model was based on a subset of patients with M-MDSC-frequencies sampled within two weeks of onset of symptoms and before potential ICU admission and who had a disease severity score less than five on the day of sampling. In the event of multiple timepoints for one patient during that period, peak M-MDSC-frequency was used.

## Data Availability

The data that support the findings of this study are available from the corresponding author, ASS, upon reasonable request.

## Acknowledgments

We thank the patients and healthy volunteers who have contributed to this study. We would also like to thank medical students and hospital staff for assistance with patient sampling and collection of clinical data. This work was supported by grants from the Swedish Research Council, the Swedish Heart-Lung Foundation, the Bill and Melinda Gates Foundation, the Knut and Alice Wallenberg Foundation and Karolinska Institutet. We like to thank Drs. Deleah Pettie, Michael Murphy, Lauren Carter and Neil P. King from the Department of Biochemistry, University of Washington, Seattle, WA, USA and Institute for Protein Design, University of Washington, Seattle, WA, USA for the production of viral proteins used in the antibody assay.

## Contributions

SF-J, SV, AL, KLo, NJ, AF and AS-S planned the study. SF-J, SV, MY, AC, IB, BÖ, MJL, IS, KLe, FH performed experiments. SF-J, RF-J, BÖ, EÅ, JS, MB, NJ, AF included and sampled patients and collected clinical data. JA provided relevant anonymized patient clinical data. SF-J, SV, MY, RF-J, AC, IB and MJL analyzed data. SF-J, MY, RF-J and MJL prepared figures. SF-J and AS-S wrote the manuscript. All co-authors edited the manuscript.

